# COVID-19 Test & Trace Success Determinants: Modeling On A Network

**DOI:** 10.1101/2020.08.05.20168799

**Authors:** Ofir Reich

## Abstract

What determines the success of a COVID-19 Test & Trace policy? We use an SEIR agent-based model on a graph, with realistic epidemiological parameters. Simulating variations in certain parameters of Testing & Tracing, we find that important determinants of successful containment are: (i) the time from symptom onset until a patient is self-isolated and tested, and (ii) the share of contacts of a positive patient who are successfully traced. Comparatively less important is (iii) the time of test analysis and contact tracing. When the share of contacts successfully traced is higher, the Test & Trace Time rises somewhat in importance. These results are robust to a wide range of values for how infectious presymptomatic patients are, to the amount of asymptomatic patients, to the network degree distribution and to base epidemic growth rate. We also provide mathematical arguments for why these simulation results hold in more general settings. Since real world Test & Trace systems and policies could affect all three parameters, Symptom Onset to Test Time should be considered, alongside test turnaround time and contact tracing coverage, as a key determinant of Test & Trace success.

## Introduction and Model Specification

The strategy of Testing & Tracing to contain the spread of COVID-19 has been proposed in many countries and contexts.[7][11][14] But which factors of Testing & Tracing should be emphasized, in order to achieve the best results? We analyze three possible determinants of contact tracing success, to determine their relative influence on epidemic growth rate and successful containment.

We use the same model as our previous paper[19], a Susceptible-Exposed-Infected-Recovered (SEIR)[13] model for disease progression. We use an agent-based model, where each agent (person) is represented as a node in a graph, and infection happens between contacts, represented by graph edges. We also model testing and self-isolation (“Quarantine”), and contact tracing of positives’ graph neighbors. In all configurations of the model, we calibrate the base infection probability so that absent any intervention, the doubling time of the number of infected is about 5 days. This sets a constant benchmark when assessing different configurations, and simulates some social distancing and hygiene measures, since without these measures the observed doubling time is around 3 days.

There are two main additions to the model presented previously: asymptomatic patients, and non-instantaneous Testing & Tracing.

### Asymptomatics

A certain share of patients are asymptomatic, independent of any other attribute (number of contacts, who they were infected from, etc.). They differ from symptomatic patients in two ways:

1. They are 50% as infectious as symptomatic patients, both in the final incubation (Exposed) phase and in the Infected phase.
2. They are never tested based on symptoms, since they do not display symptoms. They can be tested through contact tracing, and if tested before becoming Recovered they test positive.

### Non-instantaneous Testing & Tracing

Test results and contact tracing are not instantaneous, but instead take a certain amount time. See section *Model mechanics* below.

**Figure 1:**
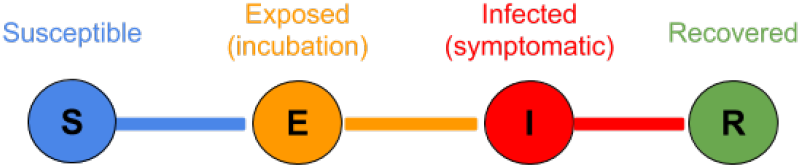
SEIR model progression for a single agent.

**Figure 2:**
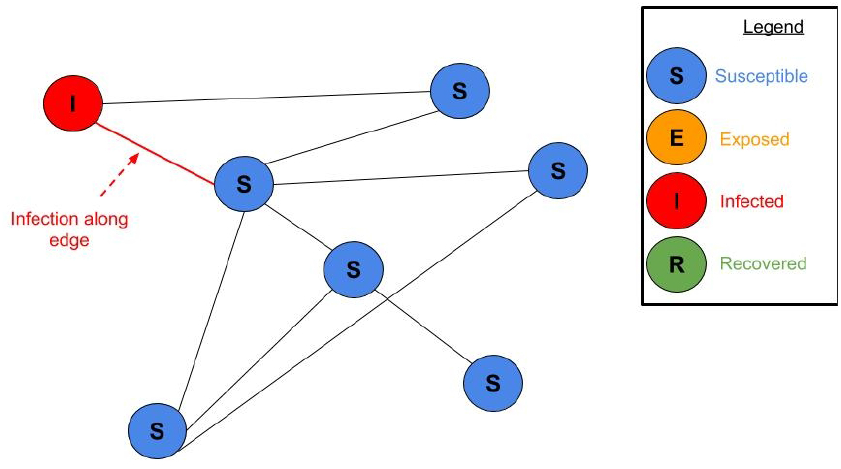
Infected nodes can infect neighboring Susceptible nodes - those who share an edge with them. Full animation at bit.ly/seir-graph-animation.

**Figure 3:**
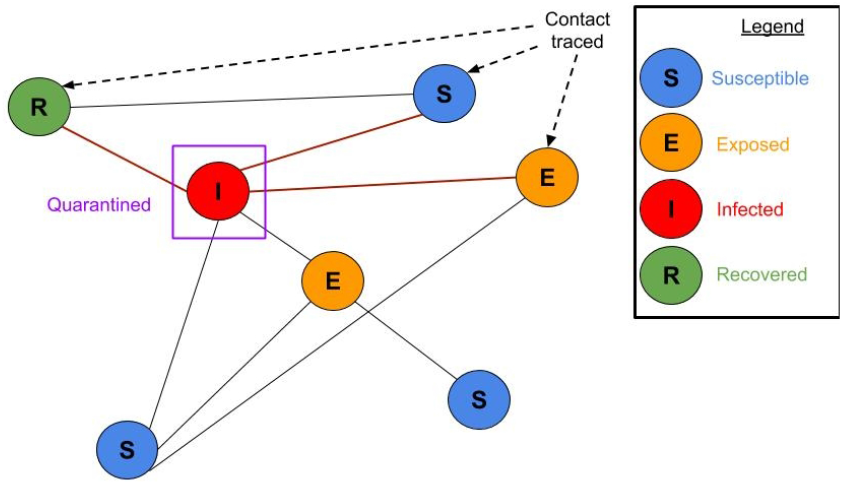
Contact tracing. ***Share of Contacts Traced*** of the neighbors of a node which tests positive are traced and themselves self-isolated and tested. In this illustration, 60% of the Infected positive node’s contacts were traced (3 out of 5).

**Figure 4:**
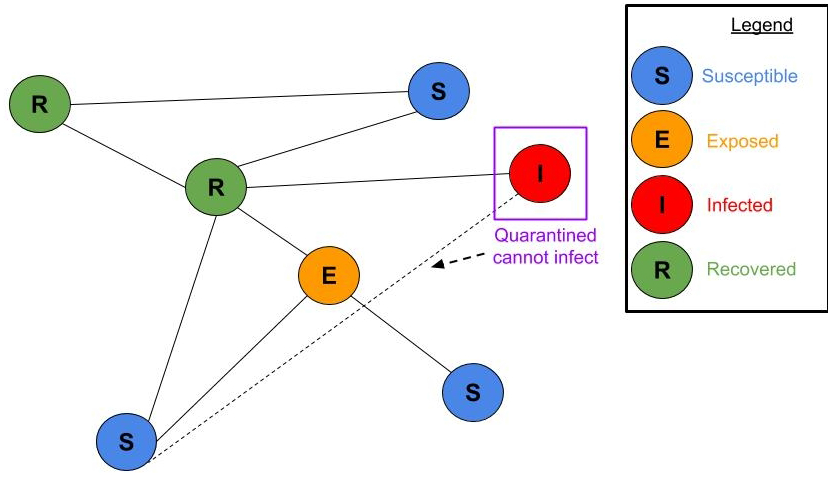
Quarantined node cannot infect others. Susceptible, Exposed, Infected and Recovered nodes shown.

**Figure 5:**
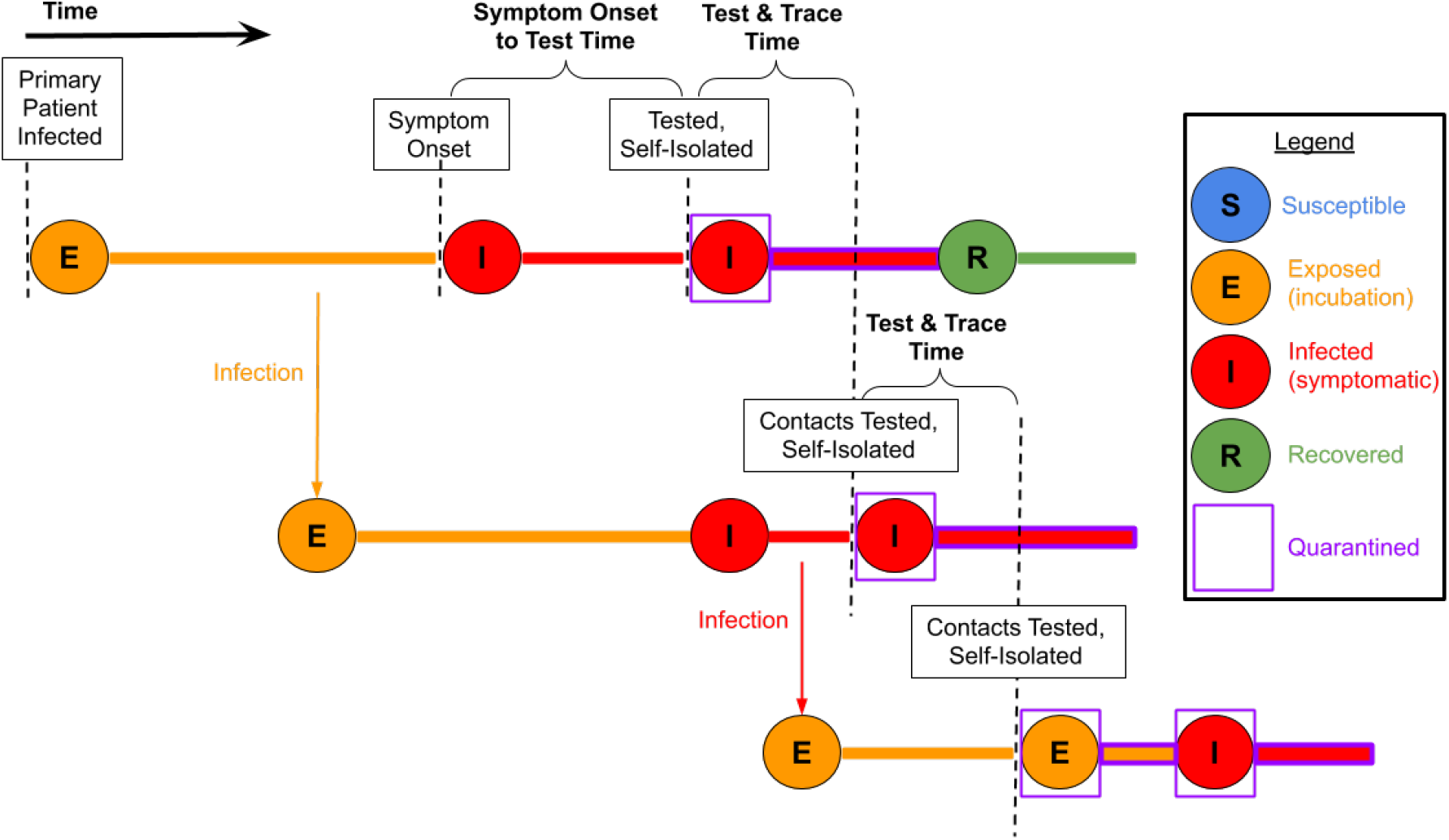
Illustration of the different time intervals in the context of disease progression. *Symptom Onset to Test Time* is the time from symptom onset to self-isolation and test. *Test & Trace Time* is the time from when the primary patient is tested and isolated until its contacts are tested and isolated. It is also the time between when the contacts are tested to when *their* contacts are isolated and tested. Not all contacts are successfully traced, only a certain fraction, *Share of Contacts Traced*.

### Model mechanics

We now describe more precisely the timeline of infection, symptoms, testing, quarantine etc. in the model. References for the values of specific parameters, where not provided here, are given in our previous paper[19]. We use the terms Quarantine, isolation and self-isolation interchangeably - they are all equivalent in our model.

1. Agent is Susceptible.
2. Agent is infected by another agent. The agent becomes Exposed, and begins its incubation phase (Gamma distributed, mean 5.1 days, std 4.4 days).
3. In the last 2 days of the incubation (Exposed) phase, it is infectious, but only 50% as much as in the Infected phase (see sensitivity analysis in presymptomatic infectiousness section).
4. Incubation ends. Agent becomes Infected, develops symptoms (for asymptomatic agents, see below).
5. After ***Symptom Onset to Test Time*** days, if the agent has not yet Recovered, it shows up to be tested. At this point it is Quarantined for 14 days, and the clock starts on test analysis.^1^ **Asymptomatic patients** - 40% of agents are asymptomatic. They have the exact same disease progression, except they are 50% as infectious as the symptomatic at each stage (see ***Caveats and limitations*** section for discussion of these numbers). They also never show up to be tested based on symptoms (but if they are tested through contact tracing, come out positive).
6. After ***Test & Trace Time*** days, the test analysis is complete and results are positive.^2^ At this point a certain fraction, ***Share of Contacts Traced***, of the positive agent’s neighbors are traced, tested and Quarantined immediately for 14 days. Therefore, we can think of ***Test & Trace Time*** as the time between a symptomatic agent being tested and their primary circle being Quarantined after a positive result.
7. Contacts who tested positive seed a new contact tracing cycle.
8. An Infected agent Recovers after (on average, Exponentially distributed) 3.5 days. It stops being infectious and symptomatic. If an agent is Recovered and has not yet been tested, it will not be tested on account of symptoms (and if tested through contact tracing will show up negative).

It is crucial that in our model isolation of a tested agent (either symptomatic or traced) happens at the moment the test is performed, not when results are back. If isolation happens only when the results are back, it is roughly equivalent to effective parameters where the test analysis time counts against both ***Symptom Onset to Test Time*** and ***Test & Trace Time***.

For more details about the model, see our previous paper[19] or the open source code on GitHub.

### Parameters of Test & Trace

We analyze three parameters of a Test & Trace policy to determine their relative importance for containment. We formally define these three parameters now.

#### Symptom Onset to Test Time

On average, how many days does it take from symptom onset until an Infected individual shows up for a test, at which point they are Quarantined even before test results are back. This is used as the expectation of an Exponential distribution sampled for each agent.

#### Test & Trace Time

(Test to Contact Test Time) - The time from the moment a primary patient is tested until its contacts (neighbors in the graph) are Quarantined and tested. This is a constant (not distributed).

#### Share of Contacts Traced

- The fraction of a positive’s neighbors which are successfully traced. Equivalently, it is the probability that each neighbor of a positive is traced. Successful tracing is sampled independently for each neighbor of a known positive, with this probability. For example, 50% means (on average) half of neighbors are traced.

**Table 1:**
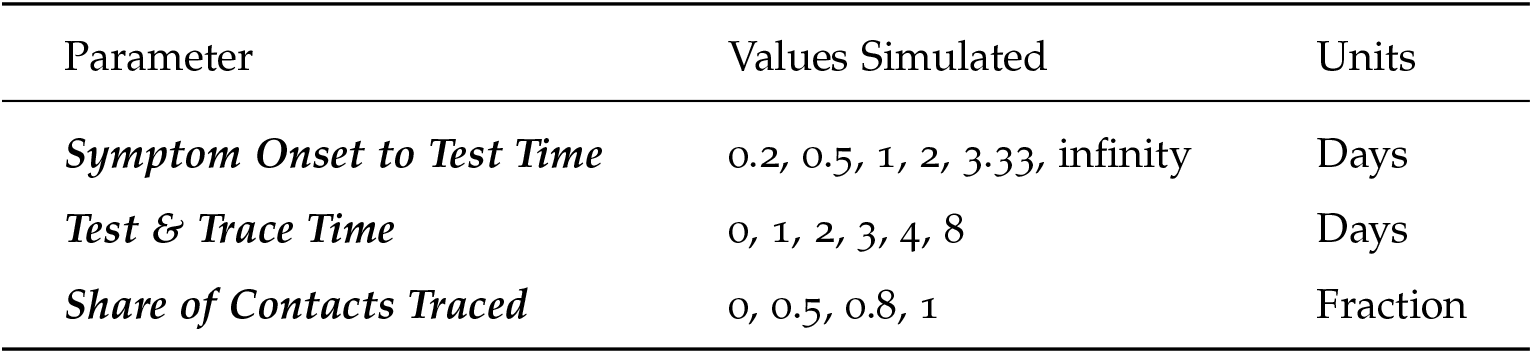
Values analyzed for the Test & Trace parameters. We simulated the cartesian product of these values.

## Simulations

### Configuration

We present simulation results. The parameters we vary in these simulations are our three analyzed parameters: ***Symptom Onset to Test Time***, ***Test & Trace Time***, and ***Share of Contacts Traced***. See the specific values used for each in Table 1 above. For each set of parameters, a full simulation of the epidemic progression on the graph is performed. The simulation runs until 600 days have been simulated, or until the epidemic is eradicated (no more Exposed or Infected nodes). Several summary statistics are collected, most prominently the fraction of the population which is ever infected, as our main outcome measure. We perform 10 independent simulations and calculate our statistics on the average of the 10 time series obtained, so each data point is the result of 10 simulations. The graph is a scalefree network of ioo,ooo nodes, with parameter gamma=0.2 (in our previous notation[19]), corresponding to a power law degree distribution with exponent −6 and mean degree 20. Each simulation is seeded with a uniformly random 0.25% of the population Infected or Exposed.

These simulations were done with some social distancing, i.e. parameters calibrated to produce a base doubling time, absent any testing and quarantine, of about 5 days. We obtained qualitatively similar results when calibrating to a doubling time of 3.1 days.

Since there is considerable uncertainty about the prevalence of asymptomatic patients, we performed a sensitivity analysis and obtained qualitatively similar results for 25% asymptomatic and for 0% asymptomatic.

We also obtained similar results for a graph which is not scalefree, but rather with a constant degree. The sensitivity analysis results are not shown here. The full raw data for these graphs is freely available.[18]

### Results

We start with an illustrative example. Figure 6 shows the trajectory of the epidemic in 4 simulations. The simulations are all with ***Share of Contacts Traced*** = 50%, but different values of ***Symptom Onset to Test Time*** and ***Test & Trace Time***, with their sum held roughly constant. Conditional on this sum, a shorter ***Symptom Onset to Test Time*** facilitates faster containment. The effect is large - the Share of population eventually infected ranges from 9.6% to 0.9%. This illustrates the greater importance of ***Symptom Onset to Test Time*** over ***Test & Trace Time***, comparing day for day.

**Figure 6:**
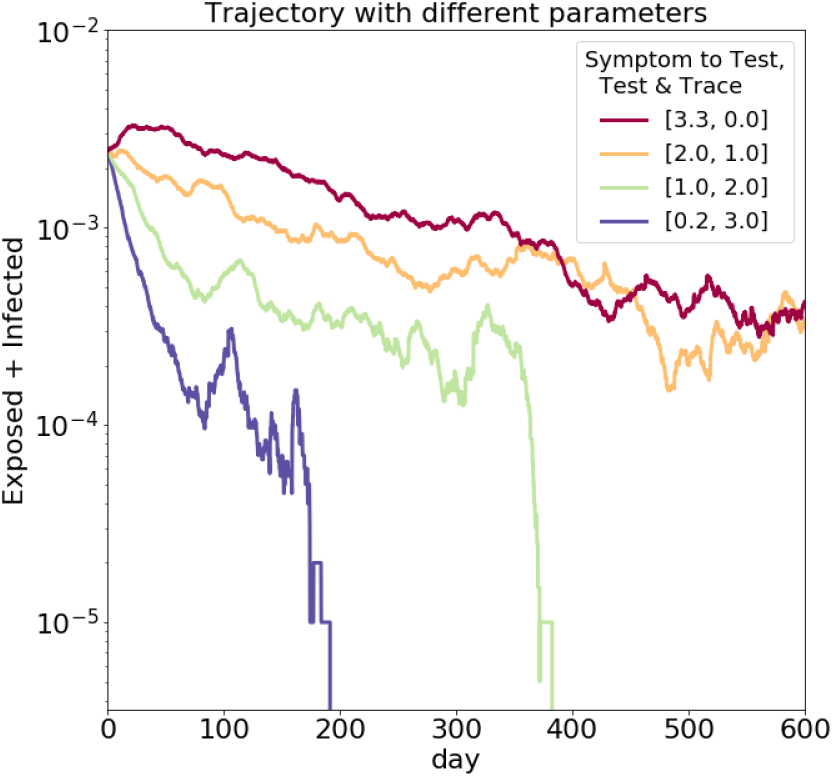
Epidemic trajectories with different time interval parameter values. Plotting over time the (log-scale) share of Exposed and Infected agents in the population. Keeping the sum ***Symptom Onset to Test Time*** + ***Test & Trace Time*** roughly constant at about 3 days, a shorter ***Symptom Onset to Test Time*** facilitates faster containment.

In what follows we abstract from the specific trajectories, and use only the share of population eventually infected as our outcome measure, so each trajectory such as the ones above will be represented by a single data point.

Figure 7 displays results of several simulations. In all of these, ***Share of Contacts Traced*** = 50%, whereas ***Symptom Onset to Test Time*** and of ***Test & Trace Time*** are varied to show their effects. Each point in this graph is a simulation result. The Y value is the fraction of the population which is eventually infected until the simulation ends. Lower values mean successful suppression. The X value is the ***Test & Trace Time***, in days. The different series (colors) are simulations with different values of ***Symptom Onset to Test Time*** - lower numbers are quicker detection. “inf” stands for “infinity”, meaning symptomatic agents are never detected. Indeed, when symptomatic agents are never detected (brown line), the ***Test & Trace Time*** does not affect the outcome, since no contact tracing takes place.

It can be observed that the vertical distance between the different lines is much larger than the vertical distance between the left and right ends of each line. This means changes in ***Symptom Onset to Test Time*** are more influential than changes in ***Test & Trace Time***. Also note the scale is smaller - there is a big difference between 1 and 2 days in ***Symptom Onset to Test Time***, whereas the X axis scale is multiple days. ***Test & Trace Time*** still does matter, especially for the right parameters (e.g. 1 day ***Symptom Onset to Test Time***).

**Figure 7:**
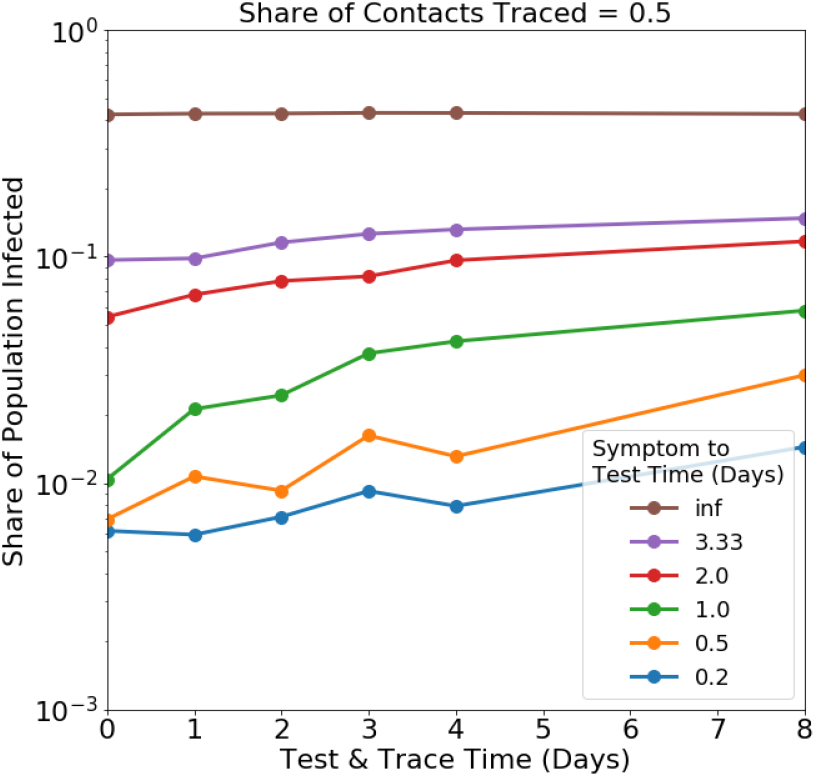
Simulation results varying ***Symptom Onset to Test Time*** and of ***Test & Trace Time***. The vertical distance between the different lines is much larger than the vertical distance between the left and right ends of each line. This means changes in ***Symptom Onset to Test Time*** are more influential than changes in ***Test & Trace Time***.

Figure 8 shows a different slice of the parameter space - we fix ***Symptom Onset to Test Time*** to 3.33 days, and show the effects of the ***Share of Contacts Traced*** (color) along with ***Test & Trace Time*** (X axis). Y is still the Share of Population Infected. As expected, when ***Share of Contacts Traced*** = 0, meaning no contacts are ever traced (red line), ***Test & Trace Time*** does not affect the outcome. The higher the share of traced neighbors (80% - brown, 100% - grey), the more influential the ***Test & Trace Time*** is. This is because the ***Test & Trace Time*** only affects the successfully traced contacts, so it is more important when more contacts are traced. Similarly, when ***Test & Trace Time*** is very long (8 days, right side of each line), the ***Share of Contacts Traced*** has a smaller effect, since contact tracing is much less effective when 8 days elapse from a positive test result to tracing of contacts - 8 days is longer than most serial intervals so the infection chain gains a longer and longer lead on the contact tracing, and not the other way around.

**Figure 8:**
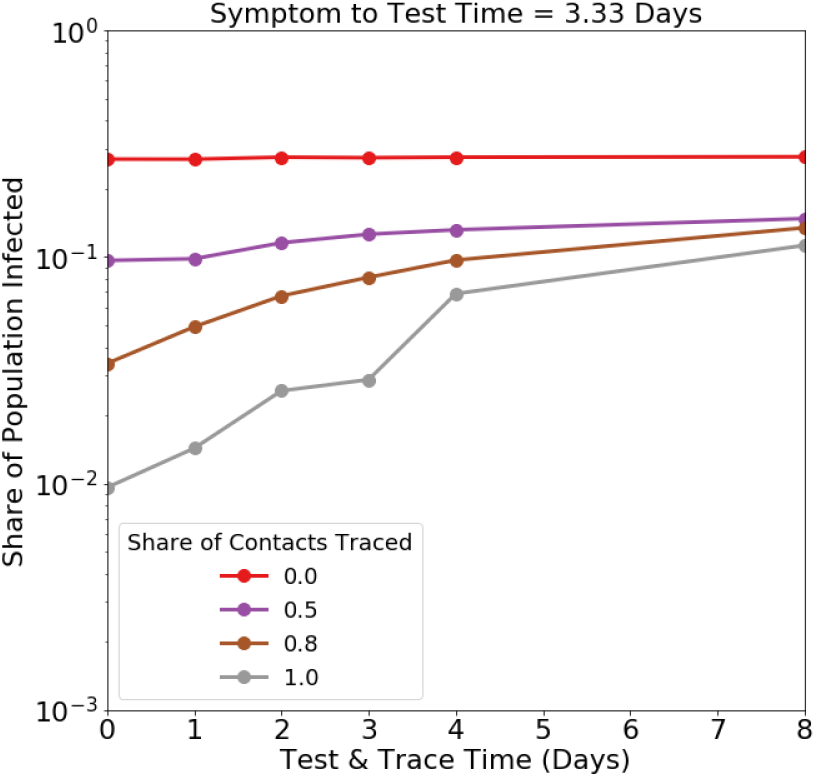
Higher ***Share of Contacts Traced*** (color) lines have a steeper slope, meaning for them ***Test & Trace Time*** (X axis) has a stronger effect.

Finally, Figure 9 shows another slice of the parameter space. We fix ***Test & Trace Time*** to 2 days, and check the relative effects of ***Symptom Onset to Test Time*** and ***Share of Contacts Traced***. Again, when no symptomatic agents are detected (***Symptom Onset to Test Time*** = infinity, brown line) the ***Share of Contacts Traced*** has no effect. But for other values, both parameters affect the Share of Population Infected - the vertical distance between the lines is comparable to the vertical distance between the right side and left side of each line.

**Figure 9:**
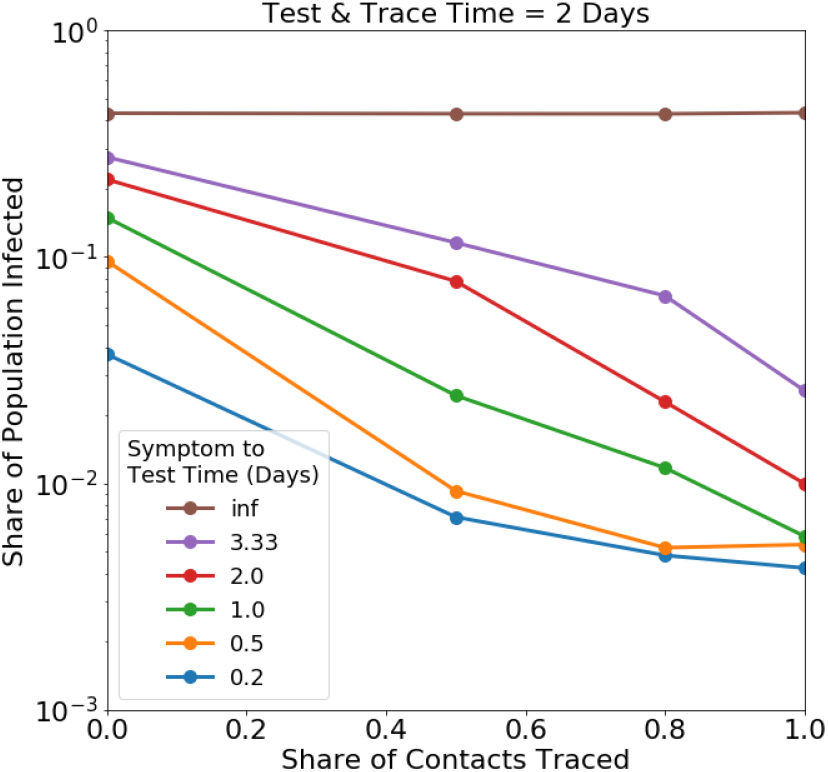
The vertical distance between the colored lines is comparable to the vertical distance between the right side and left side of each line. This means both ***Share of Contacts Traced*** and ***Symptom Onset to Test Time*** significantly affect the Share of Population Infected.

In summary, we find that ***Symptom Onset to Test Time*** and ***Share of Contacts Traced*** are stronger determinants of successful containment than ***Test & Trace Time***. For example, reducing ***Symptom Onset to Test Time*** from 2 days to 1 day has a similar effect as increasing the ***Share of Contacts Traced*** from 50% to 80%, and both are more effective than reducing the ***Test & Trace Time*** from 4 days to 1 day. When the ***Share of Contacts Traced*** is higher, ***Test & Trace Time*** rises in importance. Results for the full parameter space are presented in the first row of Figure 10.

**Figure 10:**
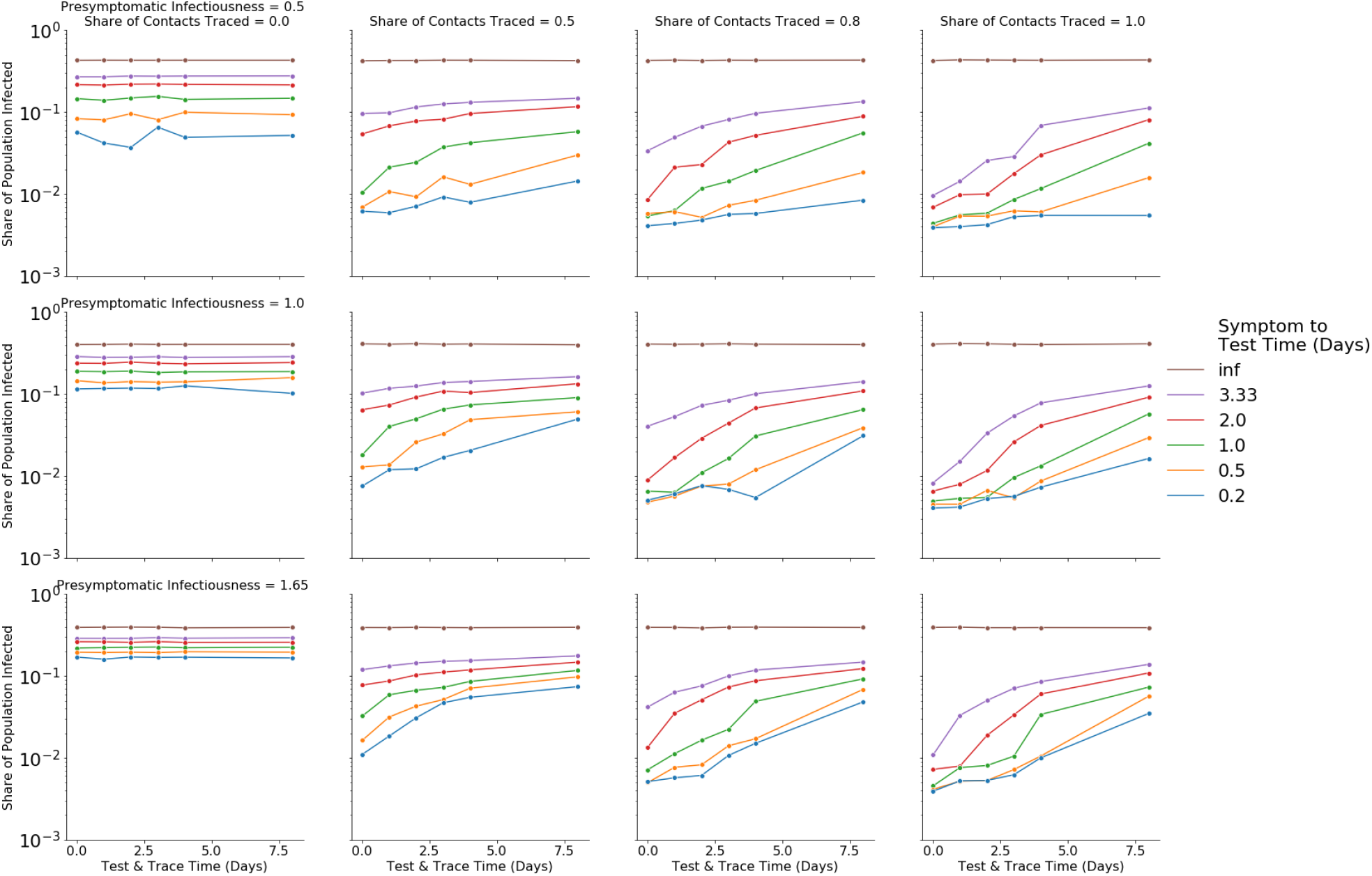
Effects of presymptomatic infectiousness. Each panel shows the effects of ***Symptom Onset to Test Time*** color) and ***Test & Trace Time*** (X axis) on Share of Population Infected (Y axis). Columns - ***Share of Contacts Traced***. Rows - presymptomatic relative infectiousness (0.5 - top, 1 - middle, 1.65 - bottom). Presymptomatic infectiousness does not change the relative importance of the main three parameters.

### Effects of presymptomatic infectiousness

We test whether greater infectiousness in the presymptomatic phase affects the results. It has been suggested[17][3][9][16] that patients are most infectious in the final days before developing symptoms, and in fact that most transmission happens before symptom onset. To simulate this, we try three different values of presymptomatic relative infectiousness. The presymptomatic relative infectiousness is the ratio between the rate at which an agent in its infectious presymptomatic phase (final 2 days of incubation) infects its neighbors, and the rate for that agent in its symptomatic phase. A value of 0.5, for example, means that if each day in the symptomatic phase, an agent infects a neighbor with probability p, then each in the presymptomatic phase that agent infects that neighbor with probability 0.5p. This parameter controls the balance between presymptomatic infections and symptomatic infections, with higher values meaning a higher share of total infections comes from the presymptomatic phase. We simulate using three values: 0.5 (the value used in results above), 1, and 1.65.^3^ For each value we recalibrate the infection probability to produce a basic doubling time (absent any intervention) of about 5 days.

Results are presented in Figure 10. Each panel is similar to 7 above, with ***Share of Contacts Traced*** fixed to a certain value, determined by the column, and with ***Symptom Onset to Test Time*** (color) and ***Test & Trace Time*** (X axis) varying. The different rows correspond to the different values of presymptomatic relative infectiousness, lower rows representing more presymptomatic infections.

Comparing the different rows, we see that increased presymptomatic infectiousness (lower rows) makes all Test & Trace efforts more difficult, which is expected since in these settings serial intervals are shorter and come before the indicator for testing - symptom onset. However, presymptomatic infectiousness does not to change the relative importance of the main three parameters - ***Share of Contacts Traced*** (columns) and ***Symptom Onset to Test Time*** (colors) still have a stronger effect than changes in ***Test & Trace Time*** (X axis value).

### Caveats and limitations

We note a few caveats and limitations of our model. Our model relies on very few unknown free parameters and magic numbers, but in return it is simplistic in many ways. We make a conscious choice not to model parameters we cannot estimate, and bear the costs in lacking representation, which is detailed here.

We model isolation of symptomatic agents and traced contacts with **perfect compliance** immediately upon test time - in reality compliance with isolation is probably higher once test results are known to be positive.

There is considerable uncertainty about the **epidemiological parameters of asymptomatic patients** - firstly their prevalence in the patient population, but also relative infectiousness and disease progression & duration. In our simulations, 40% of patients are asymptomatic[6][5][8], and have 50% relative infectiousness (see [15], and assumed by [1]). This does not contradict what we could estimate from the literature on viral shedding[20], but is still highly uncertain. Otherwise disease progression (incubation time etc.) are the same as symptomatics. Our results are robust to the share of asymptomatic patients being lower (25% or 0%, sensitivity analysis not shown), which captures a similar parameter, since the boundary between symptomatic and asymptomatic is gradual. Moreover, decreasing the infectiousness of asymptomatic patients to less than 50% would only strengthen the results.

In our model, ***Test & Trace Time* is assumed to be constant**, not randomly distributed. We don’t think this is crucial, unless test times are correlated with infectiousness (such as the degree of the node), but it could affect the results.

We use the share of the population eventually infected as our main outcome. In principle each parameter configuration results in one of two regimes: either containment, in which case the share eventually infected is proportional to the share initially infected, or mitigated spread, in which case it is proportional to the population size. In theory, the share eventually infected is an imperfect proxy for this, and the order of our results could depend on the network size or number of nodes initially infected. In practice, we observed that there is a preserved monotonic order across numbers of initial infected, so that the share eventually infected for a single number of initially infected provides a good separation between the regimes, so we do not detail it, but it is another reason not to take the absolute numbers presented as precise estimates.

All the general caveats about the model, stated in the previous paper[19], apply. For example, no modeling of false test results, an arbitrary mean degree, no graph locality, etc. Other studies have used more realistic and complex social networks[1][4] or an age structured population[2] to model epidemic spread.

## Mathematical Arguments

We provide theoretical reasons for why ***Symptom Onset to Test Time***, the time from symptom onset to test and self-isolation, might be more important for reducing growth rate than ***Test & Trace Time***. We give several reasons.

1. Reducing a day from ***Symptom Onset to Test Time***, or reducing a day from the ***Test & Trace Time***, each have the same effect on the time in which 1^st^ circle contacts are traced, tested and isolated - in both cases it makes it exactly one day earlier.
2. Reducing ***Symptom Onset to Test Time*** also reduces the amount of time the primary patient can infect, since they are isolated as soon as they are tested, not when results are back - this is not affected by ***Test & Trace Time***. In the time they are isolated, they do not infect further. So it reduces the number of additional people they infect, thereby reducing R.
3. In our model, reducing ***Symptom Onset to Test Time*** increases the share of infected agents who are ever tested, since if agents recover before their ***Symptom Onset to Test Time***, they are not tested. This gives contact tracing a better starting point - more infected agents identified. If a patient is not tested, all contacts they infected must be independently detected to break the chain.
4. ***Test & Trace Time*** only affects the ***traced*** contacts, and not the ones who aren’t traced. The effect of Test & Trace Time is therefore multiplied by the **Share of Contacts Traced**. If only 50% of contacts are traced, ***Test & Trace Time*** affects at most 50% of contacts and so only 50% of the infection chains, reducing R by at most 50%.

**Figure 11:**
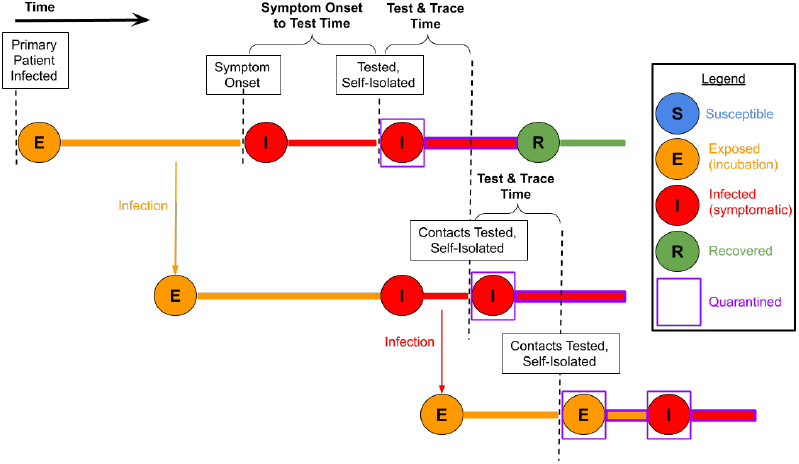
Time intervals in disease and Test & Trace progression.

## Discussion

We have found that increasing the ***Share of Contacts Traced*** and reducing ***Symptom Onset to Test Time*** are important determinants of successful contact tracing. Increasing the ***Share of Contacts Traced*** has garnered much attention[4], with proposals such as smartphone apps[10]. Contact Tracing techniques and Exposure Notifications technologies can help with all the factors we’ve examined: People who know they were exposed to a verified carrier AND develop symptoms, will get tested earlier, more contacts can be traced and of course more people will get tested as a result of getting notified about a possible exposure. We therefore focus here on the second important attribute, ***Symptom Onset to Test Time***.

How might ***Symptom Onset to Test Time*** be reduced in practice? Several policies can be considered. The first one - mentioned above, and practiced is several countries - is that self-isolation begin at the time of test sample collection (or the time of being notified of exposure, for a traced contact), not at the time when test results are back. Isolating patients only when test results are back is roughly equivalent to including the test analysis time in both the effective ***Symptom Onset to Test Time*** and the effective ***Test & Trace Time***. Symptomatic patients who are tested can be reminded or ordered to self-isolate until test results come back. Another policy is public service announcements to encourage self-isolation and testing at the first sign of symptoms, thus encouraging a quicker response. A third policy is reducing the effort and cost required to get tested, such as via drive-in testing, home testing or guidance and support hotlines (depending on test availability).

In order to track the current performance of efforts to reduce the time from symptoms to self-isolation, that time interval must be measured. It is possible to record the symptom onset time as well as the time of self-isolation and test using self-reports by patients with symptoms at sample collection sites, or patients who are contacted as part of contact tracing. For example, if a traced contact has experienced symptoms for 2 days before being contacted and has not independently asked for a test so far, we know their ***Symptom Onset to Test Time*** is greater than 2 days. With the Kaplan-Meier estimator[12], we can use these data points to estimate the distribution of the ***Symptom Onset to Test Time*** for either the entire population of traced individuals, or only the subset who end up testing positive.

So far we have considered the effects of different policies, but not the costs involved with them. We now make a few comments on cost and feasibility.

Reducing ***Symptom Onset to Test Time*** might be easier than reducing the ***Test & Trace Time***, since it involves ***one*** symptomatic patient, and not multiple contacts. It is also easier since for the typical symptomatic patient to self-isolate does not require competing resources, unlike testing and tracing capacity which need to be strengthened or diverted from other patients to speed up the Test & Trace process.

However, there are factors which make it more difficult to reduce ***Symptom Onset to Test Time***. During an advanced containment phase, the non-COVID symptomatics (due to the flu and other reasons) far outnumber the COVID symptomatics, so efforts have to impact many people who have a very weak incentive to comply. In addition, the Testing & Tracing apparatus is typically centrally controlled by the government, which makes it more amenable to tracking and improvement, as opposed to symptomatic individuals who are not in constant touch with any central agency and are much harder to monitor.

To conclude, we feel that the ***Symptom Onset to Test Time*** has been too absent from discussions about COVID-19 containment, and was not prioritized highly enough in policy.

## Data Availability

Simulation results are freely available, referenced in the body of the paper.

https://docs.google.com/spreadsheets/d/1Y65aY9FaMROYp7KT7BPJP4NqXSiNNqHZo_buQUW7yk0/edit?usp=sharing

http://www.github.com/ofir-reich/seir-graph

## Footnotes

1 If it *has* already recovered before its *Symptom Onset to Test Time*, it is not tested due to symptoms.

2 We also make the simplifying assumption that while awaiting test results, an agent is not tested again.

3 We selected 1.65 to make total presymptomatic infections roughly equal to total symptomatic infections.

## Notes

### Competing Interest Statement

The authors have declared no competing interest.

### Funding Statement

No funding was received to support this research.

### Author Declarations

Approved for publication by Google, no human subjects or data, so no IRB required.

## References

[1] Alberto Aleta, David Martin-Corral, Ana Pastore y Piontti, Marco Ajelli, Maria Litvinova, Matteo Chinazzi, Natalie E Dean, M. Elizabeth Halloran, Ira M Longini, Stefano Merler, Alex Pentland, Alessandro Vespignani, Esteban Moro, and Yamir Moreno. Modeling the impact of social distancing, testing, contact tracing and household quarantine on second-wave scenarios of the COVID-19 epidemic. May 2020. doi: 10.1101/2020.05.06.20092841. URL https://doi.org/10.1101/2020.05.06.20092841.

[2] Marie Charpignon et al. Bryan Wilder. The role of age distribution and family structure on COVID-19 dynamics: A preliminary modeling assessment for hubei and lombardy. SSRN Electronic Journal, 2020. doi: 10.2139/ssrn.3564800. URL https://doi.org/10.2139/ssrn.3564800.

[3] Zhanwei Du, Xiaoke Xu, Ye Wu, Lin Wang, Benjamin J. Cowling, and Lauren Ancel Meyers. Serial interval of COVID-19 among publicly reported confirmed cases. Emerging Infectious Diseases, 26(6):1341-1343, June 2020. doi: 10.3201/eid2606.200357. URL https://doi.org/10.3201/eid2606.200357.

[4] Adam J Kucharski et al. Effectiveness of isolation, testing, contact tracing, and physical distancing on reducing transmission of SARS-CoV-2 in different settings: a mathematical modelling study. The Lancet Infectious Diseases, June 2020. doi: 10.1016/1473-3099(20)30457-6. URL https://doi.org/10.1016/s1473-3099(20)30457-6.

[5] Daniel F. Gudbjartsson et al. Spread of SARS-CoV-2 in the icelandic population. New England Journal of Medicine, 382(24): 2302-2315, June 2020. doi: 10.1056/nejmoa2006100. URL https://doi.org/10.1056/nejmoa2006100.

[6] Enrico Lavezzo et al. Suppression of COVID-19 outbreak in the municipality of vo, italy. April 2020. doi: 10.1101/2020.04.17.20053157. URL https://doi.org/10.1101/2020.04.17.20053157.

[7] Joel Hellewell et al. Feasibility of controlling COVID-19 outbreaks by isolation of cases and contacts. The Lancet Global Health, 8(4):e488-e496, April 2020. doi: 10.1016/s2214-109x(20)30074-7. URL https://doi.org/10.1016/s2214-109x(20)30074-7.

[8] Leah F. Moriarty et al. Public health responses to COVID-19 outbreaks on cruise ships — worldwide, february–march 2020. MMWR. Morbidity and Mortality Weekly Report, 69(12): 347-352, March 2020. doi: 10.15585/mmwr.mm6912e3. URL https://doi.org/10.15585/mmwr.mm6912e3.

[9] Xi He et al. Temporal dynamics in viral shedding and transmissibility of COVID-19. Nature Medicine, 26(5)672-675, April 2020. doi: 10.1038/41591-020-0869-5. URL https://doi.org/10.1038/s41591-020-0869-5.

[10] Luca Ferretti, Chris Wymant, Michelle Kendall, Lele Zhao, Anel Nurtay, Lucie Abeler-Dörner, Michael Parker, David Bonsall, and Christophe Fraser. Quantifying SARS-CoV-2 transmission suggests epidemic control with digital contact tracing. Science, 368(6491):eabb6936, March 2020. doi: 10.1126/science.abb6936. URL https://doi.org/10.1126/science.abb6936.

[11] Josh A Firth, Joel Hellewell, Petra Klepac, Stephen M Kissler, Adam J Kucharski, and Lewis G. Spurgin and. Combining fine-scale social contact data with epidemic modelling reveals interactions between contact tracing, quarantine, testing and physical distancing for controlling COVID-19. May 2020. doi: 10.1101/2020.05.26.20113720. URL https://doi.org/10.1101/2020.05.26.20113720.

[12] Manish Kumar Goel, Pardeep Khanna, and Jugal Kishore. Understanding survival analysis: Kaplan-meier estimate. International journal of Ayurveda research, 1(4)274, 2010.

[13] William Ogilvy Kermack and Anderson G McKendrick. A contribution to the mathematical theory of epidemics. Proceedings of the royal society of london. Series A, Containing papers of a mathematical and physical character, 115(772)700-721,1927.

[14] Linda Lacina. Who coronavirus briefing: Isolation, testing and tracing comprise the ‘backbone’ of response, 2020. URL https://www.weforum.org/agenda/2020/03/testing-tracing-backbone-who-coronavirus-wednesdays-briefing/.

[15] Ruiyun Li, Sen Pei, Bin Chen, Yimeng Song, Tao Zhang, Wan Yang, and Jeffrey Shaman. Substantial undocumented infection facilitates the rapid dissemination of novel coronavirus (SARS-CoV-2). Science, 368(6490):489-493, March 2020. doi: 10.1126/science.abb3221. URL https://doi.org/10.1126/science.abb3221.

[16] Shujuan Ma, Jiayue Zhang, Minyan Zeng, Qingping Yun, Wei Guo, Yixiang Zheng, Shi Zhao, Maggie H Wang, and Zuyao Yang. Epidemiological parameters of coronavirus disease 2019: a pooled analysis of publicly reported individual data of 1155 cases from seven countries. March 2020. doi: 10.1101/2020.03.21.20040329. URL https://doi.org/10.1101/2020.03.21.20040329.

[17] Meher K Prakash. Quantitative COVID-19 infectiousness estimate correlating with viral shedding and culturability suggests 68% pre-symptomatic transmissions. May 2020. doi: 10.1101/2020.05.07.20094789. URL https://doi.org/10.1101/2020.05.07.20094789.

[18] Ofir Reich. Simulation results of test & trace success determinants - open dataset, 2020. URL https://docs.google.com/spreadsheets/d/1Y65aY9FaMROYp7KT7BPJP4NqXSiNNqHZo_buQUW7yk0/edit?usp=sharing.

[19] Ofir Reich, Guy Shalev, and Tom Kalvari. Modeling COVID-19 on a network: super-spreaders, testing and containment. May 2020. doi: 10.1101/2020.04.30.20081828. URL https://doi.org/10.1101/2020.04.30.20081828.

[20] Rui Zhou, Furong Li, Fengjuan Chen, Huamin Liu, Jiazhen Zheng, Chunliang Lei, and Xianbo Wu. Viral dynamics in asymptomatic patients with COVID-19. International Journal of Infectious Diseases, 96:288-290, July 2020. doi: 10.1016/j.ijid.2020.05.030. URL https://doi.org/10.1016/j.ijid.2020.05.030.

